# Genetic Variants in Carbohydrate Digestive Enzyme and Transport Genes Associated with Risk of Irritable Bowel Syndrome

**DOI:** 10.1101/2023.09.20.23295800

**Authors:** Hyejeong Hong, Katharina V. Schulze, Ian E. Copeland, Manasa Atyam, Kendra Kamp, Neil A. Hanchard, John Belmont, Tamar Ringel-Kulka, Margaret Heitkemper, Robert J. Shulman

**Author notes:** Corresponding author: Hyejeong Hong University of Pennsylvania, School of Nursing Department of Biobehavioral Health Sciences Claire M. Fagin Hall 418 Curie Boulevard, Room 335 Philadelphia, PA 19104 – 4217, USA.

## Abstract

Irritable Bowel Syndrome (IBS) is characterized by abdominal pain and alterations in bowel pattern, such as constipation (IBS-C), diarrhea (IBS-D), or mixed (IBS-M). Since malabsorption of ingested carbohydrates (CHO) can cause abdominal symptoms that closely mimic those of IBS, identifying genetic mutations in CHO digestive enzymes associated with IBS symptoms is critical to ascertain IBS pathophysiology. Through candidate gene association studies, we identify several common variants in *TREH*, *SI, SLC5A1* and *SLC2A5* that are associated with IBS symptoms. By investigating rare recessive Mendelian or oligogenic inheritance patterns, we identify case-exclusive rare deleterious variation in known disease genes (*SI, LCT, ALDOB,* and *SLC5A1)* as well as candidate disease genes (*MGAM* and *SLC5A2),* providing potential evidence of monogenic or oligogenic inheritance in a subset of IBS cases. Finally, our data highlight that moderate to severe IBS-associated gastrointestinal symptoms are often observed in IBS cases carrying one or more of deleterious rare variants.

## Introduction

Irritable Bowel Syndrome (IBS) is a common disorder affecting 10-15% of the global population; both adults and children.^1–5^ IBS is characterized clinically by abdominal pain associated with a change in bowel habits and categorized by bowel symptoms: IBS-Constipation (IBS-C), IBS-Diarrhea (IBS-D), and IBS-Mixed bowel patterns (IBS-M).^3,6,7^ IBS is associated with diminished quality of life among both adults and children.^8,9^

Multiple studies have sought to determine the etiology of this complex disorder. In some cases there is evidence of altered immune function systemically as well as in the gut,^10–13^ altered gut barrier function,^13^ gut dysbiosis,^14,15^ and visceral hypersensitivity^16^ among other factors. Prior enteric infection is a risk factor for post-infectious IBS.^17,18^ Additionally, results from twin studies suggest a potential genetic and environmental influence to the expression of IBS.^19,20^

Several lines of evidence suggest that variations in digestive enzyme genes influence the risk of IBS. Genome-wide association studies (GWAS) have identified approximately 26 genetic variants associated with the risk of IBS in 25 genes inclusive of *ASB4* and *SPATA5*.^21–24^ Although previous GWAS provide a valuable first insight into candidate genetic loci of IBS, the functional effect of most genetic variants from GWAS remains largely unexplained given the polygenic nature of IBS. In addition, studies have implicated rare recessive variation in genes that cause congenital deficiencies of carbohydrate (CHO) digestive enzymes,^25,26^ but the potential role of CHO digestive enzyme deficiencies in IBS is poorly defined. There is substantial clinical overlap between symptoms of IBS and the classical presentation of individuals with congenital enzymatic deficiency of these key digestive enzymes.^25,27^ Both monoallelic (homozygous recessive) and biallelic variation (compound heterozygous *in trans*) have been reported to cause congenital deficiency in sucrase-isomaltase (*SI*),^28^ maltase-glucoamylase (*MGAM*),^29^ lactase (*LCT*),^30^ trehalase (*TREH*),^31^ Solute Carrier Family 5 Member 1 (*SLC5A1*),^32^ Solute Carrier Family 2 Member 5 (*SLC2A5*),^33^ and hereditary fructose intolerance (Aldolase, Fructose-Bisphosphate B [*ALDOB*]). Given the heritability of IBS (0-57%)^28^ and the phenotypic parallels observed in individuals with congenital enzymatic deficiencies, we hypothesized that common and rare variation within the aforementioned digestive enzymes could contribute to IBS symptoms and diagnosis in a subset of patients.^34^ We undertook targeted deep-sequencing using next-generation technologies of these seven genes in a well-phenotyped IBS cohort inclusive of 687 adult and pediatric cases and 439 non-IBS controls from three regional sites in the United States. The generated data were then interrogated to identify IBS-associated common variants through candidate gene association studies as well as rare recessive Mendelian or oligogenic inheritance that would likely result in congenital deficiency.

## Results

### Demographics and Sample Characteristics

We used genomic DNA that had been collected from saliva from 229 children with IBS and 130 healthy control (HC) children at Texas Children’s Hospital (TCH) and from blood from 458 adults with IBS and 309 HCs from the University of Washington (UW) and the University of North Carolina at Chapel Hill (UNC) (Figure 1). Our IBS sample cohort included a total of 1,126 participants (687 IBS cases and 439 HCs) with a median age of 27.0 (range, 7-83). The sample was predominantly female (74.5%, n= 839) and white (87.4%, n= 691) (Table 1). To assess whether genetic ancestry could confound our results, we conducted multidimensional scaling (MDS) using single nucleotide variant (SNV) data from the 1000 Genomes Project compared with common variation from individuals within the IBS cohort (see Methods). Aside from 60 individuals who did not report their race, our cohort included persons who self-identified as Native American or Alaskan Native (n= 3), Asian (n= 28), Black (n= 55), Hispanic or Latino (n= 103), White (n= 832), and Multiple or Other (n= 45). We found that self-reported ethnicities were highly consistent with genetic ancestry, showing 93.85% concordance (Figure 2). To account for the population stratification evident in our cohort, we included MDS components as covariates in our association tests. We did not, however, see a significant difference in genetic ancestry between cases and controls (p > 0.05, Fisher’s exact test), nor among IBS subtypes.

**Figure 1.**
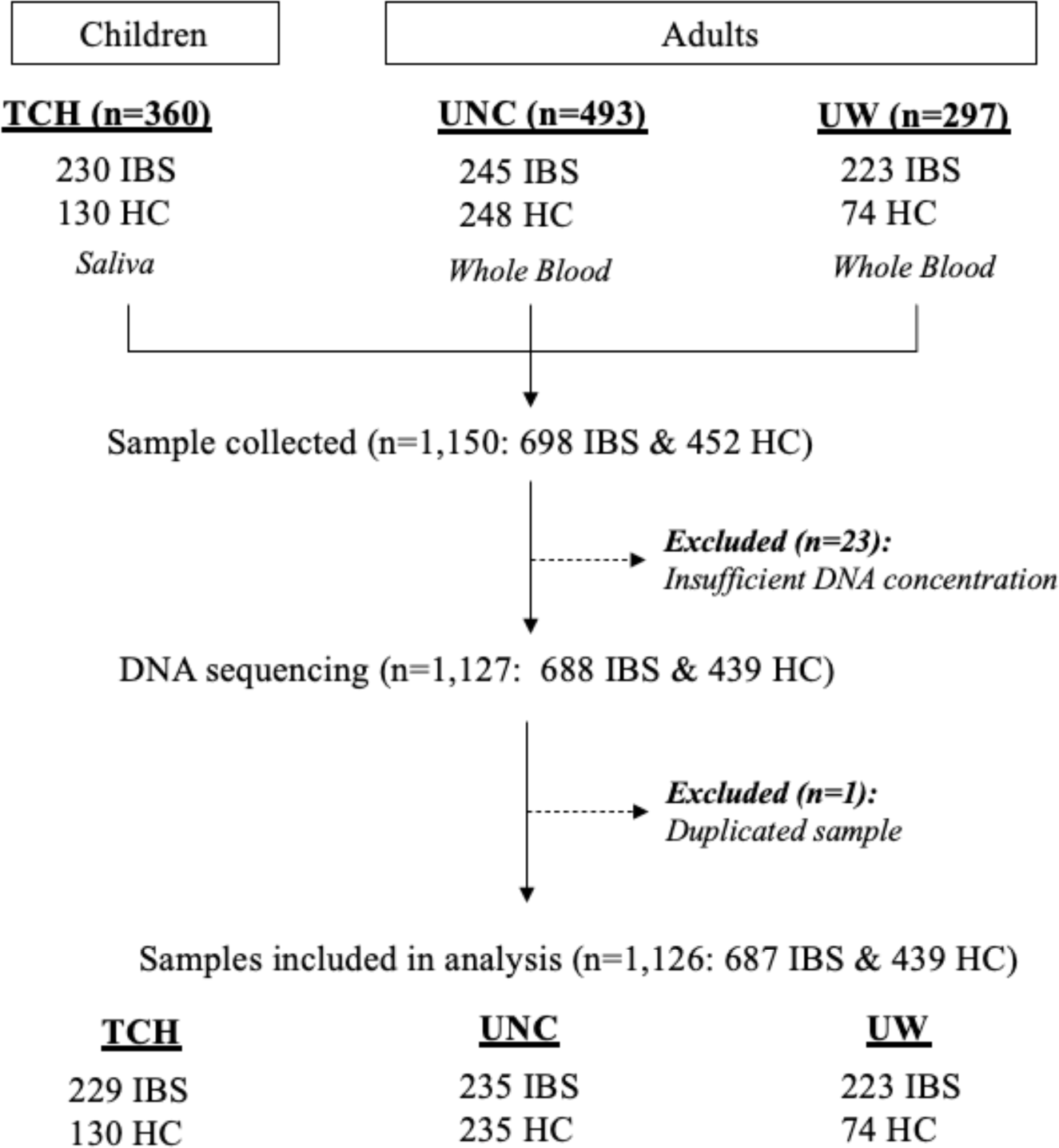
Flow of sampling from three regional study sites.

**Figure 2.**
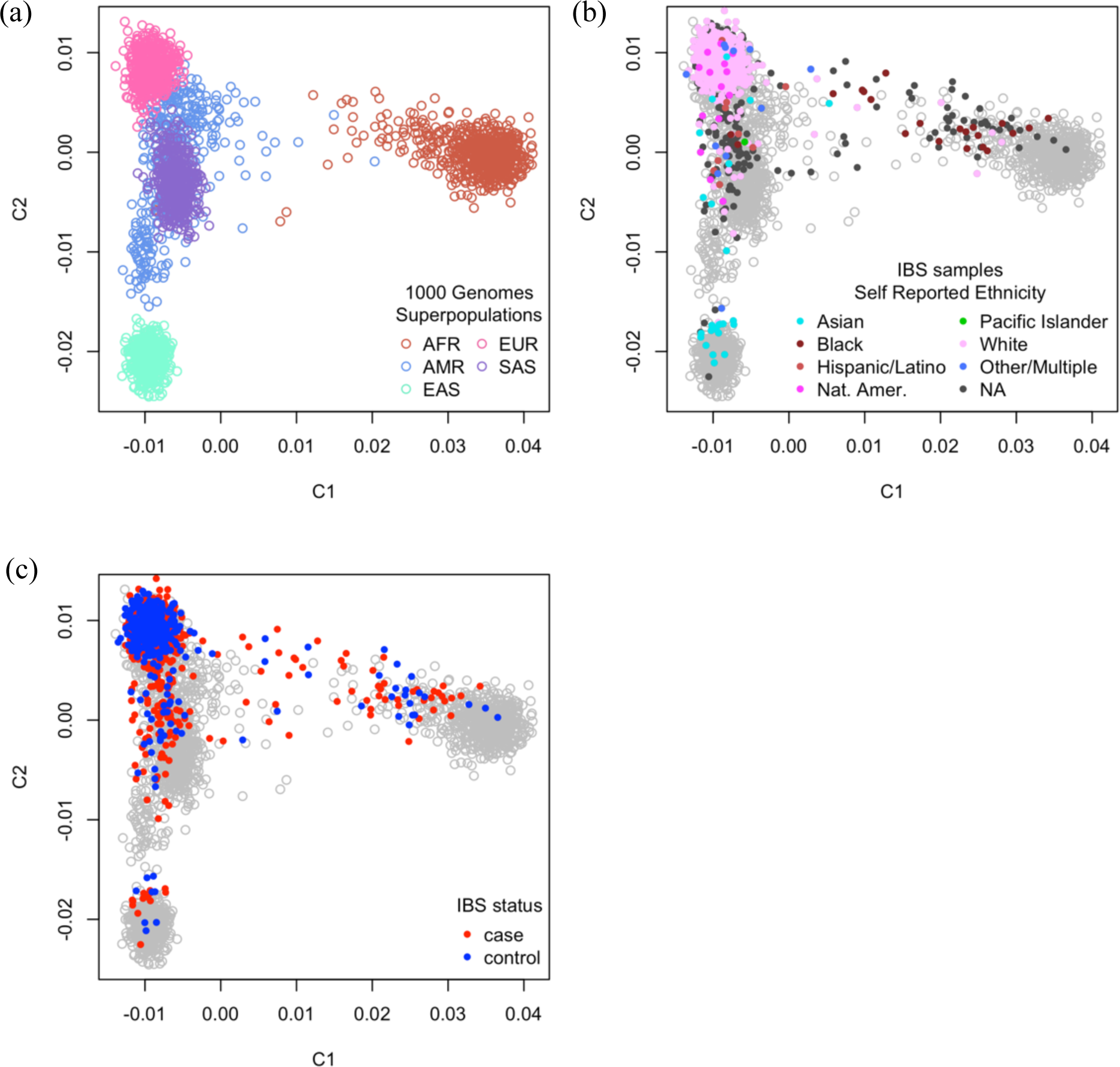
The concordance of self-reported ethnicities and genetic ancestry.

**Table 1.** Demographic characteristics of study participants.

|  | **IBS Cases**<br>N=687 | **Healthy Controls**<br>N=439 | ***p* value** |
| --- | --- | --- | --- |
| **Age – Median (IQR)**<br>N= 1,126 | 27.3 (11.0, 46.6)<br>Range: 7-81 | 26.9 (12.0, 47.0)<br>Range: 7-83 | 0.60* |
| **Female Sex – n (%)**<br>N= 1,126 | 527 (77.2) | 312 (72.1) | 0.06<sup>∞</sup> |
| **Ethnicity**<br>N= 652 |  |  | 0.60<sup>∞</sup> |
| Non-Hispanic | 377 (83.4) | 172 (85.6) |  |
| Hispanic | 74 (16.4) | 29 (14.4) |  |
| **Race**<br>N=1,066 |  |  | 0.06<sup>∞</sup> |
| Asian | 21 (3.2) | 7 (1.7) |  |
| Black | 37 (5.6) | 18 (4.4) |  |
| White | 566 (85.4) | 369 (90.7) |  |
| Other | 39 (5.9) | 13 (3.2) |  |
| **Site**<br>N=1,126 |  |  | <0.01<sup>∞</sup> |
| TCH | 229 (33.3) | 130 (29.6) |  |
| UNC | 235 (34.2) | 235 (53.5) |  |
| UW | 223 (32.5) | 74 (16.9) |  |

IQR= interquartile range; TCH= Texas Children's Hospital; UNC= University of North Carolina at Chapel Hill; UW= University of Washington
\* Correlation was evaluated using wilcoxon tests; <sup>∞</sup> Correlation was evaluated using chi-square tests.

### Common variants in CHO digestive enzyme and transport enzyme genes are associated with IBS risk

#### TREH

Association testing was conducted on IBS cases versus HCs for each candidate gene with an additive genetic model adjusted for sex and the first two MDS components. Five SNVs within *TREH* were associated with an IBS clinical diagnosis (Table 2). Of those, an increased number of minor alleles at rs45472704 and rs45529131 was positively associated with the IBS clinical phenotype with adjusted odds ratios (aORs) of 2.41 (95% confidence interval [CI]= 1.26-4.60) and 2.36 (95% CI= 1.23-4.52), respectively. The number of minor alleles at rs2277296 and rs2277297 were negatively associated with the IBS phenotype (aOR= 0.76, 95% CI= 0.62-0.93); these two SNVs were in strong linkage disequilibrium (LD, r^2^= 1), suggesting a single effect with dependent variants. Via GTEx portal, we further found that both rs2277296 and rs2277297 impact *TREH* expression levels in tissues of gastrointestinal (GI) tract. Each is an expression quantitative trait locus (eQTL) where the minor allele is associated with lower *TREH* expression in tissues, including the terminal ileum and transverse colon (Supplemental Figure 1). These two loci also showed strong evidence for regulation of transcription in the GI tract and are associated with regulatory motif alteration with Calcyclin-binding protein (*CACYBP*), according to RegulomeDB and HaploReg v4.1.^35,36^ In IBS subtype analyses, we found that the number of minor alleles at eight loci (rs17748, rs7928371, rs2276065, rs10790256, rs12225548, rs10892251, rs2277296, and rs2277297) in *TREH* are, cumulatively, significantly associated with decreased risk of IBS-C (aORs= 0.57-0.59, 95% CIs from [0.38-0.84] to [0.40-0.87]) (Table 2). Finally, the associations between genotypes and IBS-associated phenotypes were externally validated using the United Kingdom BioBank (UKBB) dataset (Supplemental Table 1a). From an independent panel from UKBB data, we also found that rs10892251, rs2277296, and rs2277297 were significantly associated with decreased sugar consumption behavior (i.e., less likely to have sugar added to tea) in the European population and metadata (β= -0.01, p< 0.01; Supplemental Table 1b).

**Table 2.**
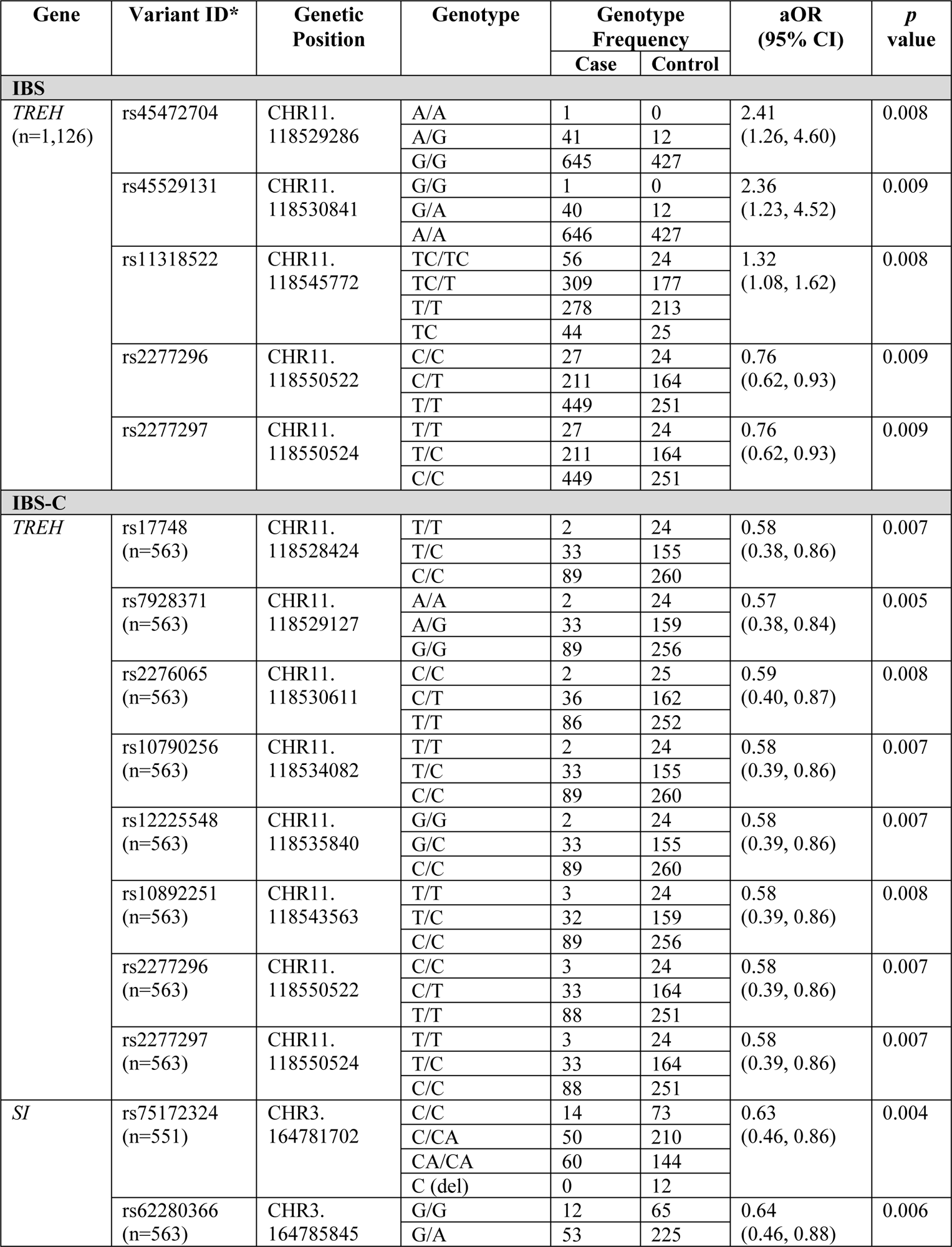
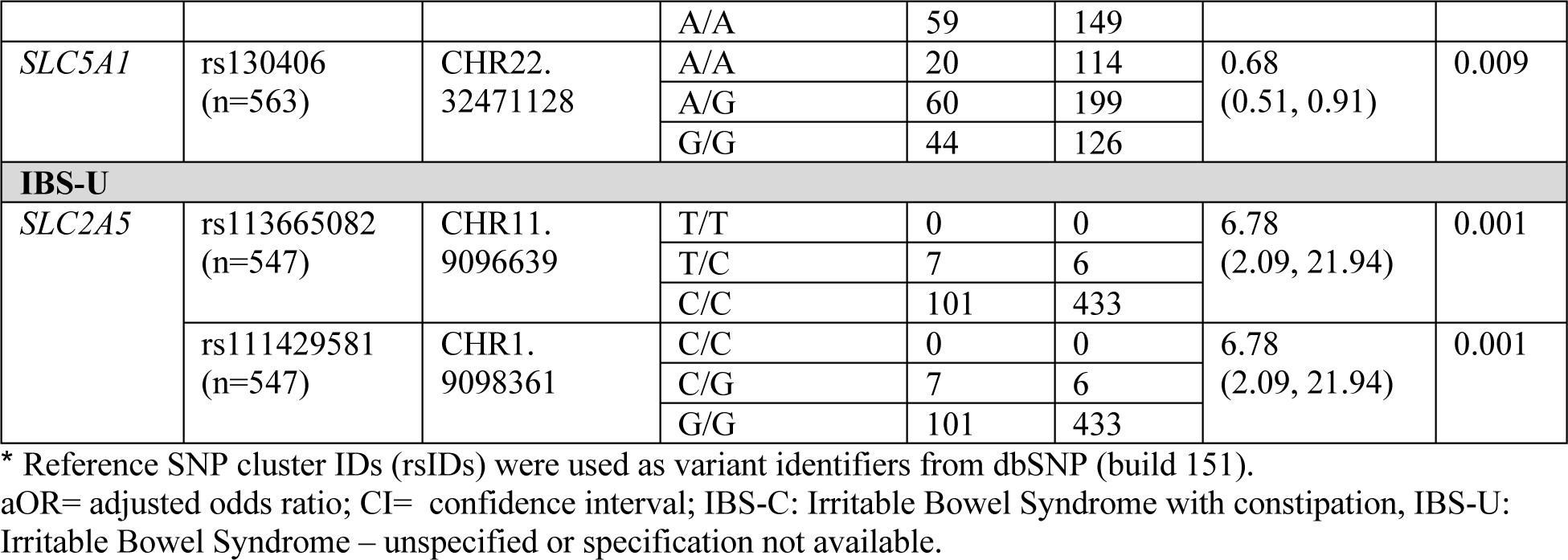
Common variants associated with IBS clinical phenotypes.

| Gene | Variant ID* | Genetic Position | Genotype | Genotype Frequency |  | aOR (95% CI) | p value |
| --- | --- | --- | --- | --- | --- | --- | --- |
|  |  |  |  | Case | Control |  |  |
| IBS |  |  |  |  |  |  |  |
| TREH<br>(n=1,126) | rs45472704 | CHR11.<br>118529286 | A/A | 1 | 0 | 2.41<br>(1.26, 4.60) | 0.008 |
|  |  |  | A/G | 41 | 12 |  |  |
|  |  |  | G/G | 645 | 427 |  |  |
|  | rs45529131 | CHR11.<br>118530841 | G/G | 1 | 0 | 2.36<br>(1.23, 4.52) | 0.009 |
|  |  |  | G/A | 40 | 12 |  |  |
|  |  |  | A/A | 646 | 427 |  |  |
|  | rs11318522 | CHR11.<br>118545772 | TC/TC | 56 | 24 | 1.32<br>(1.08, 1.62) | 0.008 |
|  |  |  | TC/T | 309 | 177 |  |  |
|  |  |  | T/T | 278 | 213 |  |  |
|  |  |  | TC | 44 | 25 |  |  |
|  | rs2277296 | CHR11.<br>118550522 | C/C | 27 | 24 | 0.76<br>(0.62, 0.93) | 0.009 |
|  |  |  | C/T | 211 | 164 |  |  |
|  |  |  | T/T | 449 | 251 |  |  |
|  | rs2277297 | CHR11.<br>118550524 | T/T | 27 | 24 | 0.76<br>(0.62, 0.93) | 0.009 |
|  |  |  | T/C | 211 | 164 |  |  |
|  |  |  | C/C | 449 | 251 |  |  |
| IBS-C |  |  |  |  |  |  |  |
| TREH | rs17748<br>(n=563) | CHR11.<br>118528424 | T/T | 2 | 24 | 0.58<br>(0.38, 0.86) | 0.007 |
|  |  |  | T/C | 33 | 155 |  |  |
|  |  |  | C/C | 89 | 260 |  |  |
|  | rs7928371<br>(n=563) | CHR11.<br>118529127 | A/A | 2 | 24 | 0.57<br>(0.38, 0.84) | 0.005 |
|  |  |  | A/G | 33 | 159 |  |  |
|  |  |  | G/G | 89 | 256 |  |  |
|  | rs2276065<br>(n=563) | CHR11.<br>118530611 | C/C | 2 | 25 | 0.59<br>(0.40, 0.87) | 0.008 |
|  |  |  | C/T | 36 | 162 |  |  |
|  |  |  | T/T | 86 | 252 |  |  |
|  | rs10790256<br>(n=563) | CHR11.<br>118534082 | T/T | 2 | 24 | 0.58<br>(0.39, 0.86) | 0.007 |
|  |  |  | T/C | 33 | 155 |  |  |
|  |  |  | C/C | 89 | 260 |  |  |
|  | rs12225548<br>(n=563) | CHR11.<br>118535840 | G/G | 2 | 24 | 0.58<br>(0.39, 0.86) | 0.007 |
|  |  |  | G/C | 33 | 155 |  |  |
|  |  |  | C/C | 89 | 260 |  |  |
|  | rs10892251<br>(n=563) | CHR11.<br>118543563 | T/T | 3 | 24 | 0.58<br>(0.39, 0.86) | 0.008 |
|  |  |  | T/C | 32 | 159 |  |  |
|  |  |  | C/C | 89 | 256 |  |  |
|  | rs2277296<br>(n=563) | CHR11.<br>118550522 | C/C | 3 | 24 | 0.58<br>(0.39, 0.86) | 0.007 |
|  |  |  | C/T | 33 | 164 |  |  |
|  |  |  | T/T | 88 | 251 |  |  |
|  | rs2277297<br>(n=563) | CHR11.<br>118550524 | T/T | 3 | 24 | 0.58<br>(0.39, 0.86) | 0.007 |
|  |  |  | T/C | 33 | 164 |  |  |
|  |  |  | C/C | 88 | 251 |  |  |
| SI | rs75172324<br>(n=551) | CHR3.<br>164781702 | C/C | 14 | 73 | 0.63<br>(0.46, 0.86) | 0.004 |
|  |  |  | C/CA | 50 | 210 |  |  |
|  |  |  | CA/CA | 60 | 144 |  |  |
|  |  |  | C (del) | 0 | 12 |  |  |
|  | rs62280366<br>(n=563) | CHR3.<br>164785845 | G/G | 12 | 65 | 0.64<br>(0.46, 0.88) | 0.006 |
|  |  |  | G/A | 53 | 225 |  |  |
|  |  |  | A/A | 59 | 149 |  |  |
| SLC5A1 | rs130406<br>(n=563) | CHR22.<br>32471128 | A/A | 20 | 114 | 0.68<br>(0.51, 0.91) | 0.009 |
|  |  |  | A/G | 60 | 199 |  |  |
|  |  |  | G/G | 44 | 126 |  |  |
| IBS-U |  |  |  |  |  |  |  |
| SLC2A5 | rs113665082<br>(n=547) | CHR11.<br>9096639 | T/T | 0 | 0 | 6.78<br>(2.09, 21.94) | 0.001 |
|  |  |  | T/C | 7 | 6 |  |  |
|  |  |  | C/C | 101 | 433 |  |  |
|  | rs111429581<br>(n=547) | CHR1.<br>9098361 | C/C | 0 | 0 | 6.78<br>(2.09, 21.94) | 0.001 |
|  |  |  | C/G | 7 | 6 |  |  |
|  |  |  | G/G | 101 | 433 |  |  |

\* Reference SNP cluster IDs (rsIDs) were used as variant identifiers from dbSNP (build 151).
aOR= adjusted odds ratio; CI= confidence interval; IBS-C: Irritable Bowel Syndrome with constipation, IBS-U: Irritable Bowel Syndrome – unspecified or specification not available.

#### SI

We observed two variants in the *SI* gene (rs75172324 and rs62280366) in 124 cases and 439 controls. The number of minor alleles of each locus had negative associations with the odds of IBS-C subtype (aOR= 0.63, 95% CI= 0.46-0.86 and aOR= 0.64, 95% CI= 0.46-0.88, respectively), suggesting that those with these variants may have a lower likelihood of constipation but potentially higher odds of diarrhea. Likewise, these two variants are also significantly associated with the increased risk of diarrhea (β= 0.15, p <0.01; Supplemental Table 1c) in the UKBB European population.

#### SLC5A1

*SLC5A1* rs130406 was observed in 124 cases and 439 controls, and the number of minor alleles was associated with the reduced odds of IBS-C (aOR= 0.68, 95% CI= 0.51-0.91), adjusted for sex and first six MDS components. In addition, rs130406 was associated with IBS in the UKBB European-GWAS (β= 0.05, p= 0.005; Supplemental Table 1d).

#### SLC2A5

*SLC2A5* rs113665082 and rs111429581 were also found to be in strong LD (r^2^=1) and were identified in 108 cases and 439 controls from the subset analysis. Given that IBS-U is defined as an unspecified subtype due to a lack of available specification data, we observed strong evidence of an association with IBS-U risk, with an aOR of 6.8 (95% CI=2.09-21.94) after controlling for sex and the first six MDS components.

### Rare, predicted deleterious variation is found exclusively among IBS cases

Rare deleterious variants were identified by assessing a deleterious scoring matrix combining damage and conservation prediction scores as well as potential monogenic or oligogenic inheritance. We further investigated the relationship between rare deleterious variants and the severity of IBS symptoms, including abdominal pain, diarrhea, constipation, and bloating based on Rome questionnaires rated on a 0-4 scale (Supplemental Table 2), subsequently rescaled to a 0-10 Likert-like scale. A single IBS case (RS16038_S63_L002) had biallelic *ALDOB* rare variants both predicted to affect splicing (NM_000035.4:c.799+6G>A and NM_000035.4:c.324+8C>G) with Transcript-inferred Pathogenicity (TRaP) scores of 0.134 and 0.249, respectively. Both have consensus benign variant classifications in ClinVar with respect to classical Hereditary Fructose Intolerance. This affected individual reported severe constipation with a score of 8 out of 10. No controls had *ALDOB* candidate rare variants. One IBS case (B-184_S147_L004) had biallelic rare missense variants in *LCT* (p.Lys88Asn and p.Arg1463Lys). Neither are present in the public databases used in this study and both are predicted to be damaging using the Combined Annotation-Dependent Depletion (CADD) scores (21.2 and 10.8, respectively), but are less clear with the rare exome variant ensemble learner (REVEL). This individual tended to have alternating diarrhea and constipation with severity scores of 8 and 6, respectively. No controls had similar candidate variants in *LCT*. We also observed one IBS case (RS2165_S355_L008) with biallelic variants in *SLC5A1* (NM_000343.4:c.1666-5T>C, TRaP score 0.127, p.Pro269His, CADD score 24.3, and REVEL score 0.59). This individual reported severe diarrhea (10 out of 10). No rare biallelic deleterious *SLC5A1* variants were observed in HCs. Rare and damaging biallelic missense variants were identified in four cases in *SI*, while no similar variants were observed among HCs (Table 3 and Supplemental Table 3). These four IBS cases with *SI* variants tended to present with moderate to severe abdominal pain, diarrhea and bloating (as rated on a scale of 6-10, Table 3).

**Table 3.**
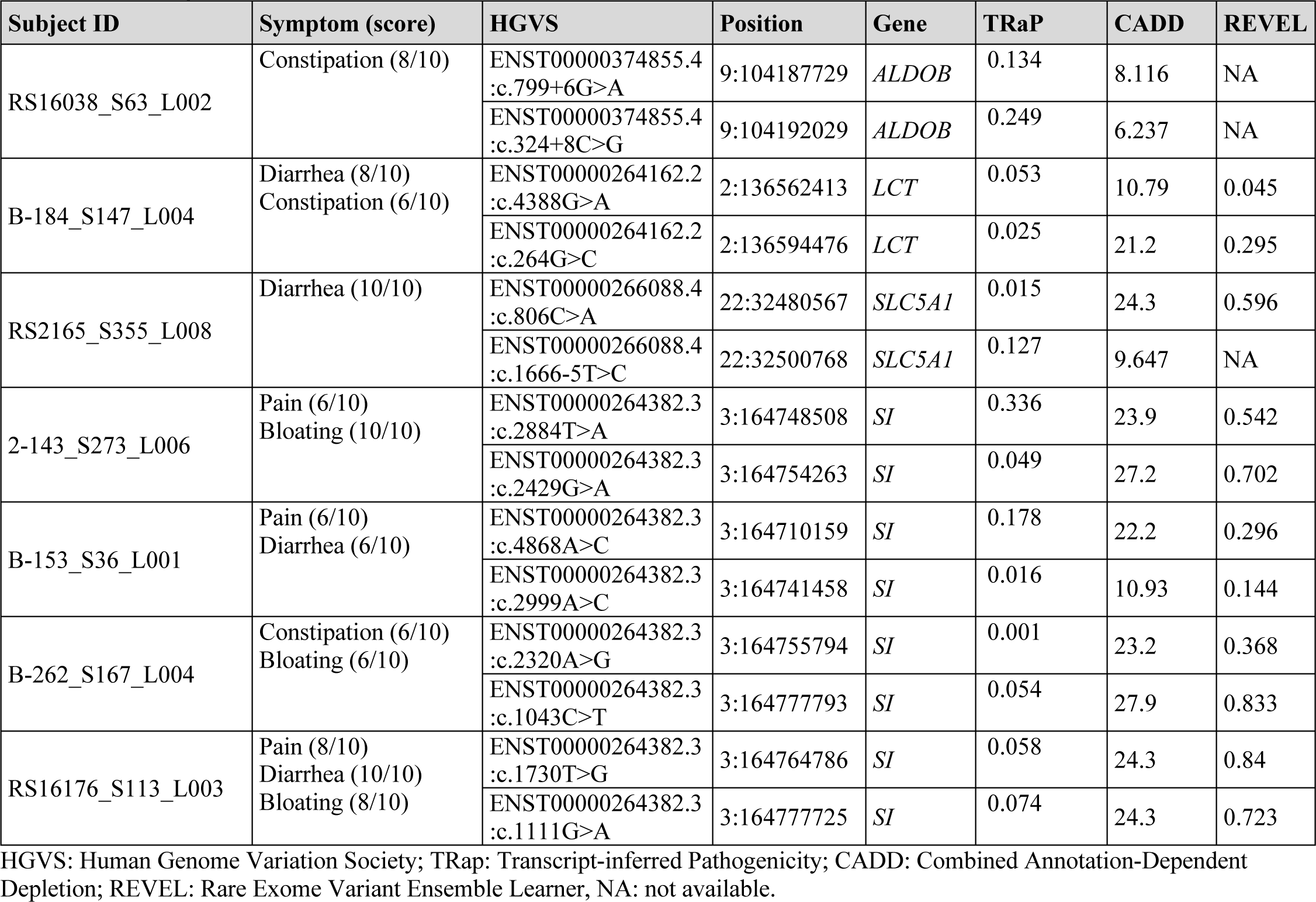
Monogenic inheritance of deleterious rare variants in IBS cases.

Using damage prediction score and conservation prediction score matrices, we identified rare and damaging biallelic missense variants in three IBS cases, and two of these three cases (RS16176_S113_L003 and 2-143_S273_L006) were identical to the variants independently identified by CADD and REVEL scores. We further validated that three variants were well conserved and predicted to be damaging to the resulting protein in ClinVar: p.Val577Gly was classified as pathogenic, p.Val371Met as likely benign, and p.Tyr975His as having conflicting interpretations (Table 4 and Supplemental Table 4). In these three IBS cases with *SI* variants, moderate to severe levels of abdominal pain and bloating were commonly reported (scored from 6 to 10). In addition, we identified rare damaging biallelic variants in *MGAM* in two IBS cases, and one of these missense variants (p.Thr1628Met) was classified as benign in ClinVar (Table 4 and Supplemental Table 4) and both cases reported moderate constipation (6 out of 10). None of the HCs had *MGAM* candidate rare variants. In none of these cases were the variants detected by genetic testing sufficient to give a diagnosis of an inherited enzyme deficiency.

**Table 4.** Rare biallelic variants determined by deleterious scoring matrix.

| Subject ID | IBS subtype | Symptom (score) | HGVS | Position | Gene | Damage Score | Conservation Score | ClinVar |
| --- | --- | --- | --- | --- | --- | --- | --- | --- |
| 2-143_S273_L006 | IBS-C | Pain (6/10)<br>Bloating (10/10) | ENST00000264382.3:c.2884T>A | 3:164748508 | *SI* | 0.83 | 0.75 | - |
|  |  |  | ENST00000264382.3:c.2429G>A | 3:164754263 | *SI* | 1 | 0.75 | - |
| RS16176_S113_L003 | IBS-D | Pain (8/10)<br>Diarrhea (10/10)<br>Bloating (8/10) | NM_001041.4:c.1730T>G | 3:164764786 | *SI* | 1 | 0.75 | Pathogenic |
|  |  |  | NM_001041.4:c.1111G>A | 3:164777725 | *SI* | 0.83 | 1 | Likely Benign |
| RS16055_S17_L001 | IBS-M | Pain (8/10)<br>Constipation (8/10)<br>Bloating (6/10) | ENST00000264382.3:c.4451G>A | 3:164716417 | *SI* | 1 | 1 | Conflicting |
|  |  |  | ENST00000264382.3:c.2923T>C | 3:164741534 | *SI* | 0.83 | 1 |  |
| RS16225_S118_L003 | IBS-U | Pain (6/10)<br>Constipation (6/10)<br>Bloating (6/10) | ENST00000549489.2:c.4652A>T | 7:141794453 | *MGAM* | 0.83 | 0.5 | - |
|  |  |  | ENST00000549489.2:c.4883C>T | 7:141795477 | *MGAM* | 0.7 | 0 | Benign |
| B-275_S192_L004 | IBS-U | Constipation (6/10) | ENST00000549489.2:c.5555C>T | 7:141805672 | *MGAM* | 0.33 | 0.25 | - |
|  |  |  | ENST00000549489.2:c.1201C>T | 7:141727515 | *MGAM* | 1 | 0.75 | - |

HGVS: Human Genome Variation Society; ClinVar: public archive of interpretations of Clinically relevant Variants; IBS-C: Irritable Bowel Syndrome with constipation, IBS-D: Irritable Bowel Syndrome with diarrhea; IBS-M: Irritable Bowel Syndrome with mixed bowel patterns; IBS-U: Irritable Bowel Syndrome – unspecified or specification not available.

We also wished to address the possibility of a cumulative intolerance to dietary carbohydrate via digenic or oligogenic inheritance. Using stringent filters for deleterious variants, we identified five cases with a rare coding or splicing variant in at least two known enzyme deficiency genes (Table 5 and Supplemental Table 5). A complex pattern of IBS symptoms was reported in these five IBS cases, with an alternating diarrhea and constipation bowel pattern being the main symptoms in three cases. None of HCs had these combinations of variants. Considering *SLC5A2* and *MGAM* as candidate disease genes, another nine cases had at least one variant from a known disease gene and these two candidate genes; while four HCs also had such combinations (Supplemental Table 5). Importantly, we identified that five IBS individuals had digenic or oligogenic recessive inheritance patterns in *SI* and *MGAM*; whereas only one HC did (Supplemental Table 5).

**Table 5.**
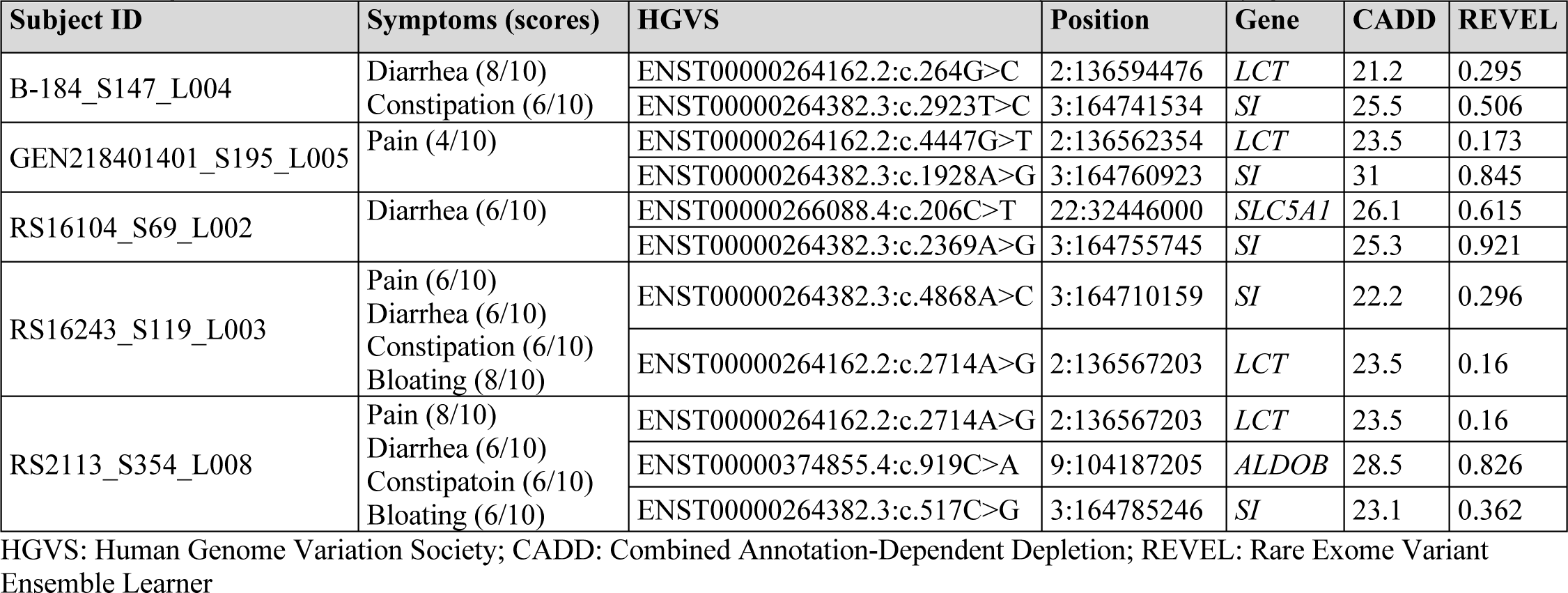
Digenic inheritance of deleterious rare variants in known enzyme deficiency genes in IBS cases.

### There was no significant rare variant burden in sequenced digestive enzyme and transport genes

To assess the involvement of rare variant burden in IBS pathophysiology, we employed the kernel-based multiple regression model to identify rare variation associated with IBS case-control phenotypes; however, the burden of rare variants (minor allele frequency [MAF] ≤ 0.05) in *ALDOB*, *TREH*, *LCT*, *MGAM*, *SI*, *SLC2A5*, and *SLC5A1* was not associated with IBS case-control phenotype (Table 6).

**Table 6.** Kernel-regression for rare variant burden analysis.

| Gene | Including all rare variants |  | Highly deleterious rare variants only |  |
| --- | --- | --- | --- | --- |
|  | Number of SNVs | *p* value | Number of SNVs | *p* value |
| *ALDOB* | 269 | 0.266398 | 10 | 0.123856 |
| *LCT* | 1055 | 0.330902 | 31 | 0.540735 |
| *MGAM* | 2461 | 0.453404 | 105 | 0.609603 |
| *SI* | 1739 | 0.83116 | 61 | 0.387663 |
| *SLC5A1* | 996 | 0.461769 | 18 | 0.182273 |
| *SLC2A5* | 692 | 0.174088 | 13 | 0.124022 |
| *TREH* | 415 | 0.294319 | 22 | 0.457838 |

SNV: single nucleotide variant

## Discussion

Although malabsorption, particularly of CHO, can cause symptoms (e.g., abdominal pain, bloating) that mimic those of IBS, there are few studies exploring the potential role of CHO digestive enzyme deficiencies in IBS. Here, we identify several common and rare variants in CHO digestive enzymes associated with IBS clinical phenotypes and symptoms in adults and children. Our results highlight that: (i) several common variants in *TREH*, *SI, SLC5A1* and *SLC2A5* are associated with IBS, and the IBS-C and IBS-U phenotypes; (ii) case-exclusive rare deleterious variation in known disease genes, such as *SI, LCT, ALDOB, SLC5A1* as well as candidate disease genes such as *MGAM* and *SLC5A2* provided potential evidence of monogenic or oligogenic inheritance in a subset of IBS cases; and (iii) moderate to severe IBS-associated GI symptoms were often observed in IBS cases carrying one or more of deleterious rare variants.

Traditionally, these enzyme deficiencies are considered rare, genetically inherited in a recessive fashion, and mainly the result of “loss-of-function” mutations.^37–40^ Although it is believed that congenital *SI* deficiencies cause symptoms recognized in infants or toddlers, more recent data suggest that the presentation may be more subtle in the case of the newly recognized allelic variants.^41,42^ Indeed, previous data from pediatric studies suggest that *SI* deficiency can be found in some children with recurrent abdominal pain, because enzyme deficiencies may be more common than previously recognized.^43,44^ In a previous IBS study in adults, a low CHO diet reduced abdominal pain and stooling frequency in some IBS-D patients, underscoring the potential role of mutations and variations in function producing IBS-like symptoms.^45^

*SI* exhibits a wide α-glucosidase activity profile and cooperates with *MGAM* in digesting α-1,4 linkages, the major glycosidic linkages in starchy foods.^46,47^ *SI* and *MGAM* are paralogues that both possess GH31 family catalytic domains and a Trefoil (P-type) domain.^48,49^ We identified the presence of rare *SI* deleterious variants with or without *MGAM* are more likely associated with an increased risk of IBS-associated GI symptoms, including abdominal pain, diarrhea, and bloating. In addition, two variants were previously reported as either pathogenic or benign in Clinvar. Although Clinvar classifications were not made with respect to an IBS phenotype, it is likely these variants contribute to a partial loss of function and impact digestion. Screening for patients with rare variants such as *SI* with or without *MGAM* may be beneficial for identifying subgroups of patients and providing personalized treatment; dietary reduction of sucrose and starch and/or enzyme supplementation may be particularly beneficial for such individuals.^38,50^

*TREH* converts trehalose to glucose.^51^ In one study, trehalase deficiency, an autosomal recessive trait, was associated with abdominal pain, distension, flatulence, vomiting and diarrhea after trehalose ingestion.^52^ Our study found that several common variants in *TREH* were associated with an IBS diagnosis. Importantly, the odds of having IBS-C is 42% less likely according to the number of minor alleles at either rs2277296 or rs2277297. Such associations are perhaps mediated by, in part, downregulation of *TREH* expression because rs2277296 and rs2277297 are eQTLs in diverse tissue types in the GI tract, such as esophageal mucosa, terminal ileum, colon, pancreas, and liver, as shown by GTEx. For example, individuals carrying two copies of minor alleles (C/C) at rs2277296 may have lower *TREH* expression in the small intestine and reduced trehalase activity, leading to an increased risk of having an IBS-D phenotype. Moreover, a subset of common variants in *TREH* (rs10892251, rs2277296, and rs2277297) may be evidence of dietary selection pressures within certain populations that have traditionally increased consumption of sugar-sweetened beverages. This supposition is explained partly by the UKBB data because minor alleles of these three variants were also associated with less sugar consumption.

We also examined genetic variations in *SLC5A1* and *SLC2A5* which are involved in glucose, galactose, and fructose intestinal uptake.^53–55^ Cellular uptake of glucose is driven by two glucose transport families: sodium driven glucose cotransporters (SGLT) and the facilitative glucose transporters (GLUT) encoded by the solute carrier genes *SLC5A1* and *SLC5A2*.^56,57^ Loss of SGLT results in glucose-galactose malabsorption, a rare congenital autosomal recessive disorder characterized by severe diarrhea starting at birth.^53,55^ Our common variant analysis detected that having at least one copy of a minor allele at rs130406 in *SLC5A1* was associated with 32% decreased risk of having an IBS-C phenotype. In addition, one IBS individual with digenic inheritance in *SI* and *SLC5A1* reported moderate diarrhea, while other IBS symptoms were reported as mild. Together, these data suggest that variants in *SLC5A1* increase the risk of an IBS-D phenotype.

Although there is conflicting clinical and experimental evidence on a pathophysiological role for the fructose transporter *GLUT5*, most studies highlight a role of *SLC2A5* gene as a major intestinal transporter for fructose and signaling molecule for the induction of down-stream genes encoding fructolytic and gluconeogenic enzymes.^58–62^ Studies from animal and cell line models support that fructose may positively regulate *SLC2A5/GLUT5* expression via transcriptional and posttranscriptional modification.^63–66^ However, *ex vivo* studies of human intestinal biopsies did not show significant genetic effects of *SLC2A5* on fructose intolerance.^67^ To understand the role of fructose intolerance in the context of IBS, we tested our hypothesis that genetic variation in *ALDOB* in conjunction with *SLC2A5* synergizes the association with the risk of fructose intolerance with GI symptoms, such as abdominal pain and diarrhea. Pathogenic mutations that strongly affect the function of *ALDOB,* a key enzyme for metabolizing fructose, are clinically evident in very few people.^68^ First, we identified that individuals with two common variants, rs113665082 and rs111429581 in *SLC2A5,* have an approximately seven times higher odds of having IBS-U. Additionally, we identified that one HC had digenic recessive inheritance patterns in *ALDOB* and *SLC2A5*; whereas no IBS case individuals did. Considering fructose intolerance has monogenic causes, our data shows that only one IBS case has rare recessive variant in *ALDOB* with moderate to severe constipation; whereas no HCs did. Together, due to its rarity, our data is limited to determine the clinical associations between the risk of IBS and monogenic or digenic inheritance in *ALDOB* with or without *SLC2A5*. Further studies are required to establish a role for these genetic variants in *ALDOB* and *SLC2A5* in heightening the risk of IBS by modulating fructose metabolism; if validated, they may provide potential targets for therapeutic dietary intervention.

Lactase persistence - the ability to digest milk lactose during adulthood – has a genetic origin; the frequency of lactase persistence is high in Northern European populations and decreases across East Asia and Southern Africa.^69^ In the present study, only two rare missense variants were identified in one IBS individual; while no HC contain these variants. Due to the dominance of European populations in our study, our ability to identify lactase-associated variants may be limited.

Our study has several limitations. First, we did not include all potential predispositions to an IBS phenotype - for example, diet, previous enteric infection, and/or stress.^70^ Rather, we focused on genetic variants associated with GI symptoms in IBS to increase understanding how CHO digestive capacity contributes to IBS manifestations. Second, we acknowledge that rare variants are commonly defined as a genetic variant with a MAF of < 0.01, which was the definition applied to our primary analysis. Since monogenic variants of common complex disorders like IBS can be very frequent, we applied a MAF of ≤ 0.05 in our subsequent monogenic and digenic variant association analyses. Finally, we conducted targeted genomic sequencing that focused on a panel of CHO digestive enzyme genes instead of using whole genome profiling, such as whole-genome or whole exome sequencing. Although these latter methods may create a greater opportunity to find pathogenic variants, targeted sequencing is an invaluable tool for mutation detection associated with greater sequencing depth allowing us to test our hypothesis that CHO malabsorption-related gene variants may play a role in IBS symptom generation. Furthermore, since targeted sequencing can screen a large number of samples with fast turnaround and reduced costs, it is clinically relevant in practice.

Our study has many strengths. First, we identified novel common and rare variants in CHO digestive enzyme genes and their association with IBS phenotypes. Such findings were previously unknown and have clinical significance, underlining the potential clinical value of genotyping. We anticipate that sequencing or genotyping efforts in individuals that have a family history of deficiencies in CHO digestive enzymes and/or early onset of IBS may facilitate precision medicine initiatives and impact treatment regimen. For example, our findings support evidence of close evolutionary and functional relationships in *SI* and *MGAM*, suggesting a role for disaccharide gene aggregation in IBS pathophysiology. For people with oligogenic inheritance with rare variants in *SI* and *MGAM,* avoiding specific dietary CHO may be beneficial. Second, due to the phenotypic complexity of IBS, we formed a consortium of clinical centers to diversify our study population, which improves statistical power. Data harmonization in the consortium enabled collection of greater racial and ethnic representation from three regional sites in the United States with a large sample size and identification of IBS subtype-defining SNVs. To avoid potential sources of bias, we used the same sequencing platform. Further, the IBS population was well-phenotyped using Rome II and III screening questionnaires. This helped maintain a consistent and accurate definition of the IBS group and minimized misclassification. Finally, the associations of common variants with IBS clinical phenotypes were externally validated using public databases, including UKBB, the largest population-based cohort in the United Kingdom, and GTEx portal, a large-scale consortium collecting tissue-specific gene expression data. Such cross-validated findings with a spectrum of phenotypes strengthen the clinical impact of candidate genes and variants on the increased risk of IBS. There is a need for future research to delve into the impact of these variants on CHO enzymatic function, the interplay between monogenic and polygenic factors in IBS risk, and the potential advantages of clinical trials for personalized dietary interventions to optimize patient care and therapeutic strategies.

## Methods

### Study Population and Sample Collection

This study used banked genomic DNA isolated from saliva and blood samples from well characterized cohorts of adults and children with IBS and HCs from three collaboration institutions: TCH, UW, and UNC (Figure 1). Institutional Review Board approval from each institute that included informed consent from participants (and assent from children) was obtained. For the IBS cohort, we included all participants who met the following criteria: (i) children and adults with well-phenotyped IBS (IBS-C, IBS-D, and IBS-M) based on Rome II (adults) and Rome III (children) screening criteria^7^. We excluded patients who had: bowel surgery; documented GI disorders (e.g., Crohn’s disease); a serious chronic medical condition (e.g., diabetes); chronic conditions with GI symptoms (e.g., cystic fibrosis) or celiac disease. The HC cohorts had no GI complaints. Details, including IBS study case definitions, are published for the TCH cohort ^15,71^, UW cohort,^72,73^ and UNC cohort^74^. A total of 1,126 samples were sequenced at the Laboratory for Translational Genomics at the USDA/ARS Children’s Nutrition Research Center at Baylor College of Medicine.

### Sequence Alignment and Variant Calling

Genomic DNA was sheared to 300-400 base-pair (bp) with a sonicator (Covaris, Fischer, San Diego) and the fragments purified. The sheared DNA from each sample was used to generate a paired-end library (Illumina TruSeq DNA Sample Prep Kit). After DNA quantitation and quality control (Biorad Experion), 500 ng of library products were hybridized to the custom capture bait material. A custom Nimblegen SeqCap EZ Choice library (Roche) was constructed to capture the full candidate genes, including 5’ untranslated region (UTR), all exons, introns, and 3’UTR. Seven candidate IBS genes were sequenced: *SI*, *MGAM, LCT, TREH, SLC5A1, SLC2A5 and ALDOB.* Samples were multiplexed (48 samples per lane) to facilitate high-depth coverage (mean coverage of >450X) of a total target length of 667 kilobase-pair (kb) on an Illumina Hiseq 2000 using 100 bp paired end reads.

Paired-read FASTQ files were aligned to the hg19 human reference genome using BWA-MEM (version 0.7.12)^29^ with shorter split hits marked as secondary (-M parameter). The functions SortSam, MarkDuplicates, and BuildBamIndex from the Picard toolkit (version 1.84)^30^ were used to sort aligned reads by coordinates, mark duplicate reads, and create index files for each alignment file. Realignment of reads was performed using the RealignerTargetCreator and IndelRealigner functions from the Genome Analysis Toolkit (GATK, version 4.0.5.1)^31^. To recalibrate base calls, GATK’s functions of BaseRecalibrator, AnalyzeCovariates, and PrintReads were used.

After removal of unmapped reads with samtools (v1.2)^32^ and subsequent indexing of cleaned alignment files with samtools’ index function, variants were called with GATK’s HaplotypeCaller. Following the creation of chromosome specific database files with GATK’s function GenomicsDBImport, joint genotyping was performed across all sequenced samples using the GenotypeGVCFs function from the same toolkit. Resulting chromosome specific joint variant call format (VCF) files were merged with GATK’s GatherVcfs function. GATK version 4.0.4.0 was used for all joint calling related functions (GenomicsDBImport, GenotypeGVCFs, and GatherVcfs).

Alongside variants within targeted gene regions the resulting joint-called VCF file also contained off-target variant calls, albeit at low coverage. Subsequent association tests were only run on targeted gene regions, the boundaries of which were determined by the first and last instance of 10X median coverage surrounding genes of interest across all sequenced samples. Per base coverage for each sample was calculated using bedtools’ coverage function (v2.26.0)^33^ on aligned and fully processed sequencing reads. All functions listed were run with default parameters unless otherwise stated.

### Genotype data quality control

In PLINK (v1.9)^75^, a total of 9,161 SNVs were called on seven targeted genes and filtered by Hardy-Weinberg Equilibrium (p < 1e^-6^), minor allele frequency (MAF ≥ 0.01), call rate (> 0.95), and linkage disequilibrium (LD, r^2^ < 0.2 in 50 bp windows with a 5 bp slide). Multiallelic loci were removed before merging IBS study samples and those from the 1000 Genomes Project Phase 3 based on the remaining variants’ dbSNP rsIDs (build 151), which were added with SnpSift’s annotate function (version 4.3t).^35^ Identity-by-state was calculated with PLINK *cluster constraint* (CC) parameters on the filtered 1,449 SNVs. Using MDS, we found no evidence of batch effects or any global influence of age or the other available demographic data from each cohort when plotting the first three dimensions. We further used the first three MDS components to calculate the Euclidean distance between every IBS sample and each group of individuals from the five 1000 Genomes superpopulations (‘AFR’: African, ‘AMR’: Admixed American, ‘EAS’: East Asian, ‘EUR’: European, and ‘SAS’: South Asian); an individual’s ancestral background was assigned based on the nearest superpopulation. To assess concordance, self-reported ethnicity labels were mapped to superpopulations as follows: ‘Native American / Alaska Native’ = EAS, ‘Asian’ = EAS or SAS, ‘Black’ = AFR, ‘Hispanic / Latino’ = AMR, ‘Native Hawaiian / Pacific Islander’ = EAS, and ‘White’ = EUR. Individuals who did not report their ethnicity and those who identified as “Multiple / Other” were omitted from concordance calculations.

### Common variant analysis

Variants in each of the seven genes were identified among 1,126 samples comprising of 687 IBS case and 439 HC samples. For each of the identified and filtered 1,449 SNVs, associations between genotype and clinical phenotype (IBS cases and HCs) were explored using an additive genetic regression model accounting for sex and the first two MDS components using PLINK (v1.9). Significant variants (MAF ≥ 0.01, p <0.01) were then reassessed across association tests using up to six MDS components. A subtype analysis was also performed for each of four IBS categories by comparing the allele frequencies between individuals in the subtype (IBS-D n= 206, IBS-C n=124, IBS-M n=47, IBS-U n=186) and the control samples, i.e., excluding cases in the other subtypes. QQ and Manhattan plots were generated using *qqman* in R (v1.0.136) for each analysis to determine deviation from expected. Results were filtered for SNVs that met a significance threshold of p < 0.01. We further queried the Broad Institute GTEx portal (https://gtexportal.org) to access tissue-specific gene expression data and identify SNVs that impact expression levels of these seven genes in GI tissues.

#### Examination of related phenotypes in the U.K. Biobank *(UKBB)*

The Pan-Ancestry GWAS of UKBB (https://pan.ukbb.broadinstitute.org/docs/technical-overview) is a publicly available resource of imputed common variant association scores that provides a multi-ancestry analysis of 7,221 phenotypes using a generalized mixed model association testing framework, spanning 16,119 GWASs performed using the UKBB cohort. Because there is no specific IBS phenotype that is defined in the same way as our primary research cohort, we reviewed the available UKBB phenotypes for phenotype overlap and for possible relevant endophenotypes (https://pan.ukbb.broadinstitute.org/downloads). We selected clinical phenotypes as listed in Supplemental Table 1a.

### Rare variant analysis

The frequency of each candidate variant was compared with MAF from the Genome Aggregation Database (gnomAD).^76^ A SNV was deemed to be rare if it had a MAF < 0.01 in gnomAD. An allelic-ratio threshold of 70:30 was used to as a quality control step to reduce false positive variation. We focused on non-synonymous variants.

#### Variant Annotation and Characterization

VarCards was used to functionally annotate the *in situ* likelihood of variants being protein-damaging, and included annotations from Combined Annotation-Dependent Depletion (CADD),^77^ Sorting Intolerant From Tolerant (SIFT),^78^ likelihood ratio test (LRT),^79^ Polymorphism Phenotyping v2 (Polyphen2),^80^ Functional Analysis through Hidden Markov Models (FATHMM)^81^ and Protein Variation Effect Analyzer (Provean).^82^ ‘Damaging’ variants were inferred from the following annotation criteria: “damaging”, “possibly damaging”, “probably damaging”, “disease causing”, and CADD scores > 12. Each variant was then given a final ‘damage prediction score’, representing the proportion of damage prediction algorithms that supported a ‘damaging’ designation for the variant in question; specifically a variant was considered damaging if it had a damage prediction score higher than three of six assessment criteria. Evolutionary conservation of non-synonymous sites, annotated by VarCards, was used for orthogonal assessment of ‘damaging’, using Genomic Evolutionary Rate Profiling (GERP),^83^ phylogenetic P-values (phyloP),^84^ SIte-specific PHYlogenetic analysis (SiPhy)^85^ and PHylogenetic Analysis with Space/Time models (phastcons).^86^ ‘Conserved’ variants were inferred from annotations as follows: >12 for SiPhy, >4.4 for GERP, >1.6 for phyloP, and > 0.5 for phastcons. Each variant was then given a final ‘conservation prediction score’ representing the proportion of conservation algorithms supporting strong conservation. A variant was classified as ‘conserved’ if it had a conserved prediction in at least two out of four assessment criteria. In addition, we also compared variant annotations using seqR^87^ as confirmation.

#### Assessment of Biallelic Variation

In order for biallelic variants to be considered as disease-causing candidates, variants were required to be seen together only in cases (i.e., not biallelic in control samples) and have been characterized as pathogenic and evolutionarily conserved. This was viewed as a conservative measure to reduce false positives given the lack of parental data to determine whether variants were in the *cis* or *trans* configuration. Candidate variants were also queried in Clinvar^88^–a large database of curated putative disease-causing variation.

#### Evaluation of cases for potential monogenic or oligogenic inheritance

VCF files for each gene (n= 7) and individual (n= 1,126, IBS n= 687, HC n= 439) were merged. The merged file was uploaded into an anVIL workspace (https://anvil.terra.bio/) and variants were annotated for filtering in seqr (https://seqr.broadinstitute.org/). Focusing on variants included in transcripts and predicted splice sites and filtering for variants with MAF ≤ 0.05, resulted in a list of 2,314 variants in 803 individuals. Cases were further prioritized by the occurrence of ≥ 2 non-synonymous variants. Finally, to increase potential interpretability of cases, we focused on variants which were more likely to be deleterious by filtering for CADD scores ≥ 10 and rare exome variant ensemble learner (REVEL)^89^ scores ≥ 0.3.

In exploratory work we considered the effect of known protein motifs in our seven candidate genes in the calibration of CADD and REVEL deleteriousness score filtering. Each protein sequence was annotated with its corresponding pfam domains. The resulting list of domains was then used to select sequences in all additional human proteins annotated with those pfam domains resulting in a set of selected pfam intervals corresponding to all those domains. CADD and REVEL scores for all possible variants and all variants reported in gnomAD exome database were collected. Protein domain annotations and deleteriousness scores were obtained in the UCSC Table Browser (https://genome.ucsc.edu/cgi-bin/hgTables) in the GRCh37/hg19 reference. Distribution of scores was analyzed in R.^90^

#### Rare variant burden analysis

VCF files corresponding to each gene (n= 7) including all individuals (n= 1,126) were parsed in PLINK1.9 (https://zzz.bwh.harvard.edu/plink) to produce binary input and site information files for association testing. Rare variant kernel-regression-based association tests were performed the R package SKAT.^91^ Three marker sets were used: (i) the full marker set consisting of both common and rare variants (n= 9,161 variant positions); (ii) rare variants only (n= 7,627); and (iii) rare variants with high deleteriousness scores (n= 260).

#### Association of rare genetic variation with IBS clinical phenotypes

We investigated the relationship between rare genetic variants and severity/frequency of IBS symptoms, including abdominal pain, diarrhea, constipation, and bloating. IBS phenotype data were collected based on a Rome II screening questionnaire in adults and Rome III screening questionnaire in children, with severity ratings ranging from 0 to 4 (Supplemental Table 2). Subsequently, IBS symptom severity scores were converted on a 0-10 likert-like scale with 0 being no symptom and 10 the worst symptom that could be imagined. Because the statistical power of classical single-variant-based association tests for low-frequency and rare variants is extremely low, we report absolute scores that reflect the severity/frequency of IBS symptoms identified by individuals found to have rare variants, instead of testing the effects of candidate rare variants on IBS symptom severity.

## Data Availability

In adherence to ethical and privacy standards, we are unable to deposit individual genotype data, while we remain committed to promoting research transparency within regulatory guidelines.

## Acknowledgements

We would like to acknowledge the clinical and research staff integral to the TCH, UNC and UW study sites, as well as the participation of the individual study participants and their families.

## References

1. Saps M, Seshadri R, Sztainberg M, Schaffer G, Marshall BM, Di Lorenzo C. A prospective school-based study of abdominal pain and other common somatic complaints in children. S0022-3476(08)00855-X pii;10.1016/j.jpeds.2008.09.047 doi. J Pediatr. Mar 2009;154(3):322–6. NOT IN FILE. doi:10.1016/j.jpeds.2008.09.047

2. Schwille IJ, Giel KE, Ellert U, Zipfel S, Enck P. A community-based survey of abdominal pain prevalence, characteristics, and health care use among children. Clin Gastroenterol Hepatol. Oct 2009;7(10):1062–8. doi:10.1016/j.cgh.2009.07.002

3. Hyams JS, Di Lorenzo C, Saps M, Shulman RJ, Staiano A, van Tilburg M. Functional Disorders: Children and Adolescents. Gastroenterology. Feb 15 2016;150:1456–1468. doi:10.1053/j.gastro.2016.02.015

4. Everhart JE, Ruhl CE. Burden of digestive diseases in the United States part II: lower gastrointestinal diseases. Research Support, N.I.H., Extramural Review. Gastroenterology. Mar 2009;136(3):741–54. doi:10.1053/j.gastro.2009.01.015

5. Everhart JE, Ruhl CE. Burden of digestive diseases in the United States part I: overall and upper gastrointestinal diseases. Research Support, U.S. Gov’t, P.H.S. Review. Gastroenterology. Feb 2009;136(2):376–86. doi:10.1053/j.gastro.2008.12.015

6. Drossman DA, Hasler WL. Rome IV-Functional GI Disorders: Disorders of Gut-Brain Interaction. Gastroenterology. May 2016;150(6):1257–61. doi:10.1053/j.gastro.2016.03.035

7. Longstreth GF, Thompson WG, Chey WD, Houghton LA, Mearin F, Spiller RC. Functional bowel disorders. S0016-5085(06)00512-9 pii;10.1053/j.gastro.2005.11.061 doi. Gastroenterology. Apr 2006;130(5):1480–91. NOT IN FILE. doi:10.1053/j.gastro.2005.11.061

8. Varni JW, Lane MM, Burwinkle TM, et al. Health-related quality of life in pediatric patients with irritable bowel syndrome: a comparative analysis. J Dev Behav Pediatr. Dec 2006;27(6):451–8. NOT IN FILE. doi:10.1097/00004703-200612000-00001

9. Cassar GE, Youssef GJ, Knowles S, Moulding R, Austin DW. Health-Related Quality of Life in Irritable Bowel Syndrome: A Systematic Review and Meta-analysis. Gastroenterology Nursing. 2020;43(3)

10. Vázquez-Frias R, Gutiérrez-Reyes G, Urbán-Reyes M, et al. Proinflammatory and anti-inflammatory cytokine profile in pediatric patients with irritable bowel syndrome. Revista de gastroenterologia de Mexico. Jan-Mar 2015;80(1):6–12. doi:10.1016/j.rgmx.2014.11.001

11. Ortiz-Lucas M, Saz-Peiro P, Sebastian-Domingo JJ. Irritable bowel syndrome immune hypothesis. Part one: the role of lymphocytes and mast cells. Rev Esp Enferm Dig. Nov 2010;102(11):637–47. NOT IN FILE. doi:10.4321/s1130-01082010001100004

12. Ortiz-Lucas M, Saz-Peiro P, Sebastian-Domingo JJ. Irritable bowel syndrome immune hypothesis. Part two: the role of cytokines. Rev Esp Enferm Dig. Dec 2010;102(12):711–7. NOT IN FILE. doi:10.4321/s1130-01082010001200006

13. Camilleri M, Lasch K, Zhou W. Irritable bowel syndrome: methods, mechanisms, and pathophysiology. The confluence of increased permeability, inflammation, and pain in irritable bowel syndrome. Am J Physiol Gastrointest Liver Physiol. Oct 2012;303(7):G775–85. doi:10.1152/ajpgi.00155.2012

14. Simren M, Barbara G, Flint HJ, et al. Intestinal microbiota in functional bowel disorders: a Rome foundation report. Consensus Development Conference Practice Guideline Research Support, Non-U.S. Gov’t. Gut. Jan 2013;62(1):159–76. doi:10.1136/gutjnl-2012-302167

15. Hollister EB, Oezguen N, Chumpitazi BP, et al. Leveraging Human Microbiome Features to Diagnose and Stratify Children with Irritable Bowel Syndrome. J Mol Diagn. May 2019;21(3):449–461. doi:10.1016/j.jmoldx.2019.01.006

16. Simren M, Tornblom H, Palsson OS, et al. Visceral hypersensitivity is associated with GI symptom severity in functional GI disorders: consistent findings from five different patient cohorts. Gut. Feb 2018;67(2):255–262. doi:10.1136/gutjnl-2016-312361

17. Card T, Enck P, Barbara G, et al. Post-infectious IBS: Defining its clinical features and prognosis using an internet-based survey. United European gastroenterology journal. 2018;6(8):1245–1253. doi:10.1177/2050640618779923

18. Thabane M, Simunovic M, Akhtar-Danesh N, et al. An outbreak of acute bacterial gastroenteritis is associated with an increased incidence of irritable bowel syndrome in children. Am J Gastroenterol. Apr 2010;105(4):933–9. doi:10.1038/ajg.2010.74

19. Levy RL, Jones KR, Whitehead WE, Feld SI, Talley NJ, Corey LA. Irritable bowel syhdrome in twins: heredity and social learning both contribute to etiology. Gastroenterology. 2001;121:799–804. NOT IN FILE.

20. Bengtson MB, Ronning T, Vatn MH, Harris JR. Irritable bowel syndrome in twins: genes and environment. Gut. Dec 2006;55(12):1754–9. NOT IN FILE. doi:10.1136/gut.2006.097287

21. Bonfiglio F, Zheng T, Garcia-Etxebarria K, et al. Female-Specific Association Between Variants on Chromosome 9 and Self-Reported Diagnosis of Irritable Bowel Syndrome. Gastroenterology. Jul 2018;155(1):168–179. doi:10.1053/j.gastro.2018.03.064

22. Bonfiglio F, Henström M, Nag A, et al. A GWAS meta-analysis from 5 population-based cohorts implicates ion channel genes in the pathogenesis of irritable bowel syndrome. Neurogastroenterol Motil. Sep 2018;30(9):e13358. doi:10.1111/nmo.13358

23. Ek WE, Reznichenko A, Ripke S, et al. Exploring the genetics of irritable bowel syndrome: a GWA study in the general population and replication in multinational case-control cohorts. Gut. Nov 2015;64(11):1774–82. doi:10.1136/gutjnl-2014-307997

24. Holliday EG, Attia J, Hancock S, et al. Genome-wide association study identifies two novel genomic regions in irritable bowel syndrome. Am J Gastroenterol. 2014:770–2. vol. 5.

25. Diekmann L, Pfeiffer K, Naim HY. Congenital lactose intolerance is triggered by severe mutations on both alleles of the lactase gene. BMC Gastroenterol. Mar 21 2015;15:36. doi:10.1186/s12876-015-0261-y

26. Jacob R, Zimmer KP, Schmitz J, Naim HY. Congenital sucrase-isomaltase deficiency arising from cleavage and secretion of a mutant form of the enzyme. J Clin Invest. Jul 2000;106(2):281–7. doi:10.1172/jci9677

27. Misselwitz B, Butter M, Verbeke K, Fox MR. Update on lactose malabsorption and intolerance: pathogenesis, diagnosis and clinical management. Gut. Nov 2019;68(11):2080–2091. doi:10.1136/gutjnl-2019-318404

28. Marcadier JL, Boland M, Scott CR, et al. Congenital sucrase-isomaltase deficiency: identification of a common Inuit founder mutation. Cmaj. Feb 3 2015;187(2):102–107. doi:10.1503/cmaj.140657

29. Nichols BL, Avery SE, Karnsakul W, et al. Congenital maltase-glucoamylase deficiency associated with lactase and sucrase deficiencies. J Pediatr Gastroenterol Nutr. Oct 2002;35(4):573–9. doi:10.1097/00005176-200210000-00022

30. Wanes D, Husein DM, Naim HY. Congenital Lactase Deficiency: Mutations, Functional and Biochemical Implications, and Future Perspectives. Nutrients. 2019;11(2):461. doi:10.3390/nu11020461

31. Murray IA, Coupland K, Smith JA, Ansell ID, Long RG. Intestinal trehalase activity in a UK population: establishing a normal range and the effect of disease. Br J Nutr. Mar 2000;83(3):241–5. doi:10.1017/s0007114500000313

32. Martín MG, Turk E, Kerner C, Zabel B, Wirth S, Wright EM. Prenatal identification of a heterozygous status in two fetuses at risk for glucose-galactose malabsorption. Prenat Diagn. May 1996;16(5):458–62. doi:10.1002/(sici)1097-0223(199605)16:5<458::aid-pd873>3.0.co;2-u

33. Oh A-R, Sohn S, Lee J, et al. ChREBP deficiency leads to diarrhea-predominant irritable bowel syndrome. Metabolism: clinical and experimental. 2018;85:286–297. doi:10.1016/j.metabol.2018.04.006

34. Saito YA. The role of genetics in IBS. Gastroenterology clinics of North America. 2011;40(1):45–67. doi:10.1016/j.gtc.2010.12.011

35. Boyle AP, Hong EL, Hariharan M, et al. Annotation of functional variation in personal genomes using RegulomeDB. Genome Res. Sep 2012;22(9):1790–7. doi:10.1101/gr.137323.112

36. Ward LD, Kellis M. HaploReg v4: systematic mining of putative causal variants, cell types, regulators and target genes for human complex traits and disease. Nucleic acids research. 2016;44(D1):D877–D881. doi:10.1093/nar/gkv1340

37. Husein DM, Rizk S, Naim HY. Differential Effects of Sucrase-Isomaltase Mutants on Its Trafficking and Function in Irritable Bowel Syndrome: Similarities to Congenital Sucrase-Isomaltase Deficiency. Nutrients. 2020;13(1):9. doi:10.3390/nu13010009

38. Henström M, Diekmann L, Bonfiglio F, et al. Functional variants in the sucrase-isomaltase gene associate with increased risk of irritable bowel syndrome. Gut. Feb 2018;67(2):263–270. doi:10.1136/gutjnl-2016-312456

39. Garcia-Etxebarria K, Zheng T, Bonfiglio F, et al. Increased Prevalence of Rare Sucrase-isomaltase Pathogenic Variants in Irritable Bowel Syndrome Patients. Clin Gastroenterol Hepatol. Oct 2018;16(10):1673–1676. doi:10.1016/j.cgh.2018.01.047

40. Husein DM, Naim HY. Impaired cell surface expression and digestive function of sucrase-isomaltase gene variants are associated with reduced efficacy of low FODMAPs diet in patients with IBS-D. Gut. Aug 2020;69(8):1538–1539. doi:10.1136/gutjnl-2019-319411

41. Sander P, Alfalah M, Keiser M, et al. Novel mutations in the human sucrase-isomaltase gene (SI) that cause congenital carbohydrate malabsorption. Hum Mutat. Jan 2006;27(1):119. doi:10.1002/humu.9392

42. Alfalah M, Keiser M, Leeb T, Zimmer KP, Naim HY. Compound heterozygous mutations affect protein folding and function in patients with congenital sucrase-isomaltase deficiency. S0016-5085(08)02052-0 pii;10.1053/j.gastro.2008.11.038 doi. Gastroenterology. Mar 2009;136(3):883–92. NOT IN FILE. doi:10.1053/j.gastro.2008.11.038

43. Robayo-Torres CC, Baker SS, Chumpitazi BP, Lecea CE, Nichols BL, Jr., Opekun AR. Poor starch digestion in children with CSID and recurrent abdominal pain. J Pediatr Gastroenterol Nutr. Nov 2012;55 Suppl 2:S32–4. doi:10.1097/01.mpg.0000421407.88128.5c

44. Rathod S, Friesen CA, Radford K, Colombo JM. Sucrase Breath Testing in Children Presenting With Chronic Abdominal Pain. Clin Pediatr (Phila). Nov 2020;59(13):1191–1194. doi:10.1177/0009922820942183

45. Austin GL, Dalton CB, Hu Y, et al. A very low-carbohydrate diet improves symptoms and quality of life in diarrhea-predominant irritable bowel syndrome. Clin Gastroenterol Hepatol. Jun 2009;7(6):706–708.e1. doi:10.1016/j.cgh.2009.02.023

46. Lin AH, Hamaker BR, Nichols BL, Jr. Direct starch digestion by sucrase-isomaltase and maltase-glucoamylase. J Pediatr Gastroenterol Nutr. Nov 2012;55 Suppl 2:S43–5. doi:10.1097/01.mpg.0000421414.95751.be

47. Lee BH, Eskandari R, Jones K, et al. Modulation of starch digestion for slow glucose release through “toggling” of activities of mucosal α-glucosidases. J Biol Chem. Sep 14 2012;287(38):31929–38. doi:10.1074/jbc.M112.351858

48. Naumoff DG. Structure and evolution of the mammalian maltase-glucoamylase and sucrase-isomaltase genes. Molecular Biology. 2007/12/01 2007;41(6):962–973. doi:10.1134/S0026893307060131

49. Sim L, Willemsma C, Mohan S, Naim HY, Pinto BM, Rose DR. Structural basis for substrate selectivity in human maltase-glucoamylase and sucrase-isomaltase N-terminal domains. The Journal of biological chemistry. 2010;285(23):17763–17770. doi:10.1074/jbc.M109.078980

50. Zheng T, Camargo-Tavares L, Bonfiglio F, Marques FZ, Naim HY, D’Amato M. Rare Hypomorphic Sucrase Isomaltase Variants in Relation to Irritable Bowel Syndrome Risk in UK Biobank. Gastroenterology. Nov 2021;161(5):1712–1714. doi:10.1053/j.gastro.2021.06.063

51. Murray IA, Coupland K, Smith JA, Ansell ID, Long RG. Intestinal trehalase activity in a UK population: establishing a normal range and the effect of disease. British Journal of Nutrition. 2000;83(3):241–245. doi:10.1017/S0007114500000313

52. Montalto M, Gallo A, Ojetti V, Gasbarrini A. Fructose, trehalose and sorbitol malabsorption. Eur Rev Med Pharmacol Sci. 2013;17 Suppl 2:26–9.

53. Xin B, Wang H. Multiple sequence variations in SLC5A1 gene are associated with glucose–galactose malabsorption in a large cohort of Old Order Amish. Clinical Genetics. 2011;79(1):86–91. 10.1111/j.1399-0004.2010.01440.x

54. Taneva I, Grumann D, Schmidt D, et al. Gene variants of the SLC2A5 gene encoding GLUT5, the major fructose transporter, do not contribute to clinical presentation of acquired fructose malabsorption. BMC Gastroenterol. Apr 6 2022;22(1):167. doi:10.1186/s12876-022-02244-7

55. Alamoudi LO, Alfaraidi AT, Althagafi SS, Al-Thaqafy MS, Hasosah M. Congenital Glucose-Galactose Malabsorption: A Case With a Novel SLC5A1 Mutation in a Saudi Infant. Cureus. 2021;13(10):e18440–e18440. doi:10.7759/cureus.18440

56. Deng D, Yan N. GLUT, SGLT, and SWEET: Structural and mechanistic investigations of the glucose transporters. Protein Sci. 2016;25(3):546–558. doi:10.1002/pro.2858

57. Sabino-Silva R, Mori RC, David-Silva A, Okamoto MM, Freitas HS, Machado UF. The Na(+)/glucose cotransporters: from genes to therapy. Braz J Med Biol Res. Nov 2010;43(11):1019–26. doi:10.1590/s0100-879x2010007500115

58. Patel C, Douard V, Yu S, Tharabenjasin P, Gao N, Ferraris RP. Fructose-induced increases in expression of intestinal fructolytic and gluconeogenic genes are regulated by GLUT5 and KHK. Am J Physiol Regul Integr Comp Physiol. Sep 2015;309(5):R499–509. doi:10.1152/ajpregu.00128.2015

59. Barone S, Fussell SL, Singh AK, et al. Slc2a5 (Glut5) is essential for the absorption of fructose in the intestine and generation of fructose-induced hypertension. J Biol Chem. Feb 20 2009;284(8):5056–66. doi:10.1074/jbc.M808128200

60. Ebert K, Ewers M, Bisha I, et al. Identification of essential amino acids for glucose transporter 5 (GLUT5)-mediated fructose transport. J Biol Chem. Feb 9 2018;293(6):2115–2124. doi:10.1074/jbc.RA117.001442

61. Taneva I, Grumann D, Schmidt D, et al. Gene variants of the SLC2A5 gene encoding GLUT5, the major fructose transporter, do not contribute to clinical presentation of acquired fructose malabsorption. BMC gastroenterology. 2022;22(1):167–167. doi:10.1186/s12876-022-02244-7

62. Barone S, Fussell SL, Singh AK, et al. Slc2a5 (Glut5) is essential for the absorption of fructose in the intestine and generation of fructose-induced hypertension. The Journal of biological chemistry. 2009;284(8):5056–5066. doi:10.1074/jbc.M808128200

63. Shu R, David ES, Ferraris RP. Dietary fructose enhances intestinal fructose transport and GLUT5 expression in weaning rats. Am J Physiol. Mar 1997;272(3 Pt 1):G446–53. doi:10.1152/ajpgi.1997.272.3.G446

64. Suzuki T, Douard V, Mochizuki K, Goda T, Ferraris Ronaldo P. Diet-induced epigenetic regulation in vivo of the intestinal fructose transporter Glut5 during development of rat small intestine. Biochemical Journal. 2011;435(1):43–53. doi:10.1042/bj20101987

65. Mesonero J, Matosin M, Cambier D, Rodriguez-Yoldi MJ, Brot-Laroche E. Sugar-dependent expression of the fructose transporter GLUT5 in Caco-2 cells. Biochemical Journal. 1995;312(3):757–762. doi:10.1042/bj3120757

66. Gouyon F, Onesto C, Dalet V, Pages G, Leturque A, Brot-Laroche E. Fructose modulates GLUT5 mRNA stability in differentiated Caco-2 cells: role of cAMP-signalling pathway and PABP (polyadenylated-binding protein)-interacting protein (Paip) 2. Biochemical Journal. 2003;375(1):167–174. doi:10.1042/bj20030661

67. Wilder-Smith CH, Li X, Ho SS, et al. Fructose transporters GLUT5 and GLUT2 expression in adult patients with fructose intolerance. United European Gastroenterology Journal. 2014;2(1):14–21. 10.1177/2050640613505279

68. Gibson PR, Newnham E, Barrett JS, Shepherd SJ, Muir JG. Review article: fructose malabsorption and the bigger picture. Aliment Pharmacol Ther. Feb 15 2007;25(4):349–63. doi:10.1111/j.1365-2036.2006.03186.x

69. Anguita-Ruiz A, Aguilera CM, Gil Á. Genetics of Lactose Intolerance: An Updated Review and Online Interactive World Maps of Phenotype and Genotype Frequencies. Nutrients. 2020;12(9):2689. doi:10.3390/nu12092689

70. Zia JK, Lenhart A, Yang PL, et al. Risk Factors for Abdominal Pain-Related Disorders of Gut-Brain Interaction in Adults and Children: A Systematic Review. Gastroenterology. Oct 2022;163(4):995–1023.e3. doi:10.1053/j.gastro.2022.06.028

71. Shulman RJ, Hollister EB, Cain K, et al. Psyllium Fiber Reduces Abdominal Pain in Children With Irritable Bowel Syndrome in a Randomized, Double-Blind Trial. Clin Gastroenterol Hepatol. May 2017;15(5):712–719.e4. doi:10.1016/j.cgh.2016.03.045

72. Heitkemper MM, Cain KC, Deechakawan W, et al. Anticipation of public speaking and sleep and the hypothalamic-pituitary-adrenal axis in women with irritable bowel syndrome. Neurogastroenterol Motil. Jul 2012;24(7):626–31, e270-1. doi:10.1111/j.1365-2982.2012.01915.x

73. Jarrett ME, Cain KC, Burr RL, Hertig VL, Rosen SN, Heitkemper MM. Comprehensive self-management for irritable bowel syndrome: randomized trial of in-person vs. combined in-person and telephone sessions. The American journal of gastroenterology. 2009;104(12):3004–3014. doi:10.1038/ajg.2009.479

74. Hod K, Ringel Y, van Tilburg MAL, Ringel-Kulka T. Bloating in Irritable Bowel Syndrome Is Associated with Symptoms Severity, Psychological Factors, and Comorbidities. Digestive Diseases and Sciences. 2019/05/01 2019;64(5):1288–1295. doi:10.1007/s10620-018-5352-5

75. Chang CC, Chow CC, Tellier LC, Vattikuti S, Purcell SM, Lee JJ. Second-generation PLINK: rising to the challenge of larger and richer datasets. Gigascience. 2015;4:7. doi:10.1186/s13742-015-0047-8

76. Karczewski KJ, Francioli LC, Tiao G, et al. The mutational constraint spectrum quantified from variation in 141,456 humans. Nature. 2020/05/01 2020;581(7809):434–443. doi:10.1038/s41586-020-2308-7

77. Rentzsch P, Witten D, Cooper GM, Shendure J, Kircher M. CADD: predicting the deleteriousness of variants throughout the human genome. Nucleic Acids Research. 2018;47(D1):D886–D894. doi:10.1093/nar/gky1016

78. Sim N-L, Kumar P, Hu J, Henikoff S, Schneider G, Ng PC. SIFT web server: predicting effects of amino acid substitutions on proteins. Nucleic acids research. 2012;40(Web Server issue):W452–W457. doi:10.1093/nar/gks539

79. Chun S, Fay JC. Identification of deleterious mutations within three human genomes. Genome research. 2009;19(9):1553–1561. doi:10.1101/gr.092619.109

80. Adzhubei I, Jordan DM, Sunyaev SR. Predicting functional effect of human missense mutations using PolyPhen-2. Current protocols in human genetics. 2013;Chapter 7:Unit7.20-Unit7.20. doi:10.1002/0471142905.hg0720s76

81. Shihab HA, Gough J, Cooper DN, et al. Predicting the functional, molecular, and phenotypic consequences of amino acid substitutions using hidden Markov models. Human mutation. 2013;34(1):57–65. doi:10.1002/humu.22225

82. Choi Y, Chan AP. PROVEAN web server: a tool to predict the functional effect of amino acid substitutions and indels. Bioinformatics (Oxford, England). 2015;31(16):2745–2747. doi:10.1093/bioinformatics/btv195

83. Cooper GM, Stone EA, Asimenos G, et al. Distribution and intensity of constraint in mammalian genomic sequence. Genome research. 2005;15(7):901–913. doi:10.1101/gr.3577405

84. Pollard KS, Hubisz MJ, Rosenbloom KR, Siepel A. Detection of nonneutral substitution rates on mammalian phylogenies. Genome research. 2010;20(1):110–121. doi:10.1101/gr.097857.109

85. Garber M, Guttman M, Clamp M, Zody MC, Friedman N, Xie X. Identifying novel constrained elements by exploiting biased substitution patterns. *Bioinformatics (Oxford*, England). 2009;25(12):i54–i62. doi:10.1093/bioinformatics/btp190

86. Siepel A, Bejerano G, Pedersen JS, et al. Evolutionarily conserved elements in vertebrate, insect, worm, and yeast genomes. Genome research. 2005;15(8):1034–1050. doi:10.1101/gr.3715005

87. Jadwiga Słowik MB. seqR: fast and comprehensive k-mer counting package. https://github.com/slowikj/seqR

88. Landrum MJ, Lee JM, Benson M, et al. ClinVar: improving access to variant interpretations and supporting evidence. Nucleic Acids Res. Jan 4 2018;46(D1):D1062–d1067. doi:10.1093/nar/gkx1153

89. Ioannidis NM, Rothstein JH, Pejaver V, et al. REVEL: An Ensemble Method for Predicting the Pathogenicity of Rare Missense Variants. American journal of human genetics. 2016;99(4):877–885. doi:10.1016/j.ajhg.2016.08.016

90. RStudio Team. RStudio: Integrated Development for R. RStudio, PBC, Boston, MA. http://www.rstudio.com

91. Wu MC, Lee S, Cai T, Li Y, Boehnke M, Lin X. Rare-variant association testing for sequencing data with the sequence kernel association test. American journal of human genetics. 2011;89(1):82–93. doi:10.1016/j.ajhg.2011.05.029

