## Supplemental Figure 1 for "Genetic Variants in Carbohydrate Digestive Enzyme and Transport Genes Associated with Risk of Irritable Bowel Syndrome"

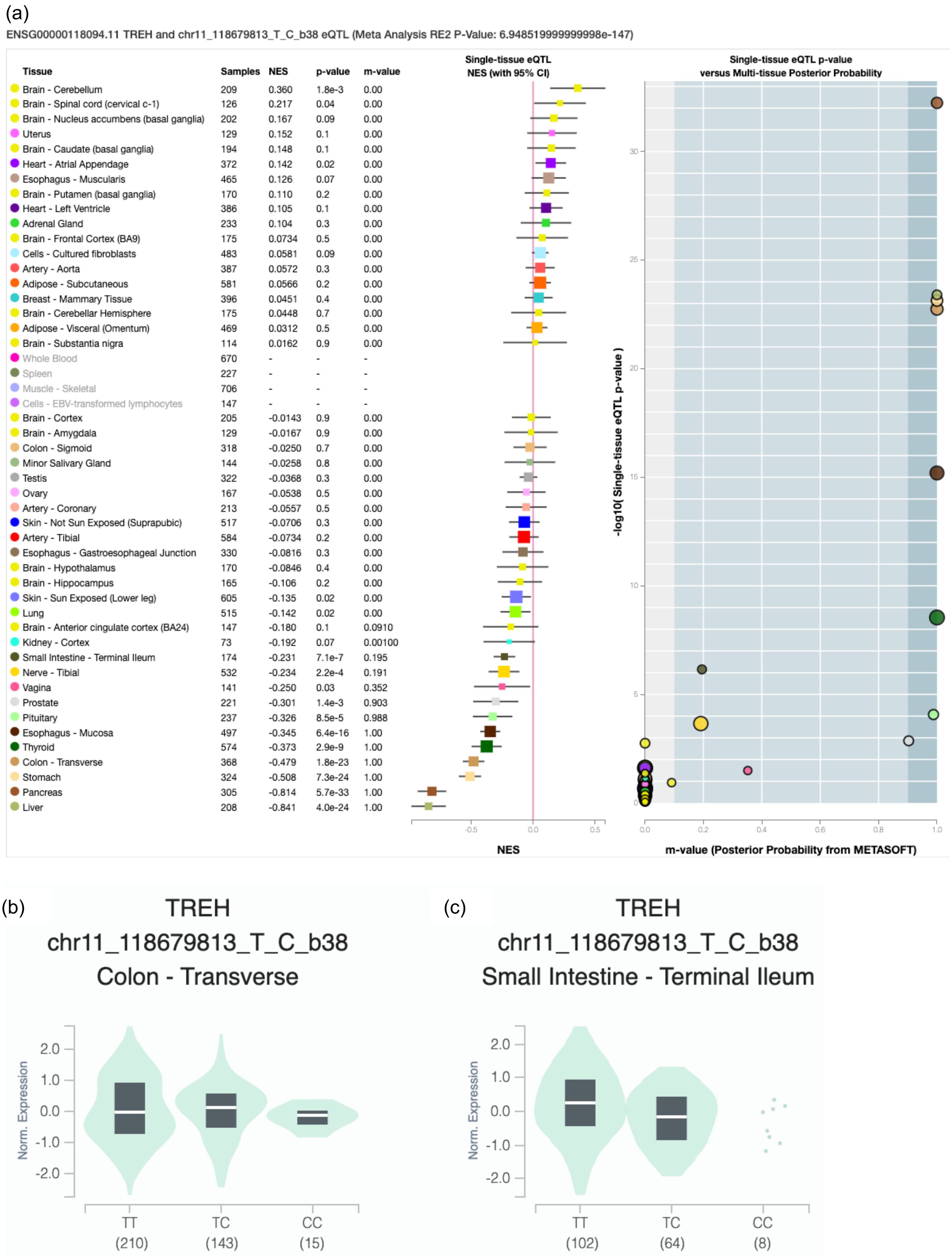
(a)
ENSG00000118094.11 TREH and chr11_118679813_T_C_b38 eQTL (Meta Analysis RE2 P-Value: 6.948519999999998e-147)
Tissue Samples NES p-value m-value
Brain - Cerebellum 209 0.360 1.8e-3 0.00
Brain - Spinal cord (cervical c-1) 126 0.217 0.04 0.00
Brain - Nucleus accumbens (basal ganglia) 202 0.167 0.09 0.00
Uterus 129 0.152 0.1 0.00
Brain - Caudate (basal ganglia) 194 0.148 0.1 0.00
Heart - Atrial Appendage 372 0.142 0.02 0.00
Esophagus - Muscularis 465 0.126 0.07 0.00
Brain - Putamen (basal ganglia) 170 0.110 0.2 0.00
Heart - Left Ventricle 386 0.105 0.1 0.00
Adrenal Gland 233 0.104 0.3 0.00
Brain - Frontal Cortex (BA9) 175 0.0734 0.5 0.00
Cells - Cultured fibroblasts 483 0.0581 0.09 0.00
Artery - Aorta 387 0.0572 0.3 0.00
Adipose - Subcutaneous 581 0.0566 0.2 0.00
Breast - Mammary Tissue 396 0.0451 0.4 0.00
Brain - Cerebellar Hemisphere 175 0.0448 0.7 0.00
Adipose - Visceral (Omentum) 469 0.0312 0.5 0.00
Brain - Substantia nigra 114 0.0162 0.9 0.00
Whole Blood 670 - - -
Spleen 227 - - -
Muscle - Skeletal 706 - - -
Cells - EBV-transformed lymphocytes 147 - - -
Brain - Cortex 205 -0.0143 0.9 0.00
Brain - Amygdala 129 -0.0167 0.9 0.00
Colon - Sigmoid 318 -0.0250 0.7 0.00
Minor Salivary Gland 144 -0.0258 0.8 0.00
Testis 322 -0.0368 0.3 0.00
Ovary 167 -0.0538 0.5 0.00
Artery - Coronary 213 -0.0557 0.5 0.00
Skin - Not Sun Exposed (Suprapubic) 517 -0.0706 0.3 0.00
Artery - Tibial 584 -0.0734 0.2 0.00
Esophagus - Gastroesophageal Junction 330 -0.0816 0.3 0.00
Brain - Hypothalamus 170 -0.0846 0.4 0.00
Brain - Hippocampus 165 -0.106 0.2 0.00
Skin - Sun Exposed (Lower leg) 605 -0.135 0.02 0.00
Lung 515 -0.142 0.02 0.00
Brain - Anterior cingulate cortex (BA24) 147 -0.180 0.1 0.0910
Kidney - Cortex 73 -0.192 0.07 0.00100
Small Intestine - Terminal Ileum 174 -0.231 7.1e-7 0.195
Nerve - Tibial 532 -0.234 2.2e-4 0.191
Vagina 141 -0.250 0.03 0.352
Prostate 221 -0.301 1.4e-3 0.903
Pituitary 237 -0.326 8.5e-5 0.988
Esophagus - Mucosa 497 -0.345 6.4e-16 1.00
Thyroid 574 -0.373 2.9e-9 1.00
Colon - Transverse 368 -0.479 1.8e-23 1.00
Stomach 324 -0.508 7.3e-24 1.00
Pancreas 305 -0.814 5.7e-33 1.00
Liver 208 -0.841 4.0e-24 1.00
Single-tissue eQTL NES (with 95% CI)
-0.5
0.0
0.5
NES
Single-tissue eQTL p-value versus Multi-tissue Posterior Probability
-log10( Single-tissue eQTL p-value )
0
5
10
15
20
25
30
0.0
0.2
0.4
0.6
0.8
1.0
m-value (Posterior Probability from METASOFT)
(b)
TREH
chr11_118679813_T_C_b38
Colon - Transverse
Norm. Expression
2.0
1.0
0.0
-1.0
-2.0
TT (210)
TC (143)
CC (15)
(c)
TREH
chr11_118679813_T_C_b38
Small Intestine - Terminal Ileum
Norm. Expression
2.0
1.0
0.0
-1.0
-2.0
TT (102)
TC (64)
CC (8)

**Supplemental Figure 1. Identification of eQTLs for rs2277296 in Multiple GI Tissues from GTEx.** [Top] (a) In the forest plot, the error bars represent the 95% confidence intervals (standard errors) of the normalized effect sizes (NES, equivalent to the beta coefficient) of rs2277296 on the expression of *TREH* for specific tissue types. It reveals a positive association with several brain tissues and a negative association with tissues in multiple systems, including the digestive system (small intestine, colon, stomach, pancreas), the nervous system (tibial nerve, pituitary), the reproductive system (vagina, prostate), as well as skin and lung. [Bottom] In the violin plots, the x-axis represents the genotype of rs2277296 and shows the number of GTEx samples with that genotype listed in parentheses, while the y-axis represents normalized *TREH* gene expression levels. The minor allele "C" of rs2277296 is associated with decreased expression of *TREH* (b) in transverse colon tissues (NES = -0.48, p = $1.8e^{-23}$) and (c) in terminal ileum tissues of the small intestine (NES = -0.23, p = $7.07e^{-7}$).

Note. Only the eQTL plots for rs2277296 are displayed in this figure since the eQTL plots for rs2277296 and rs2277297 are identical due to their high linkage disequilibrium (LD, $r^2$=1).

*We acknowledge the Genotype-Tissue Expression (GTEx) Project, which received support from the Common Fund of the Office of the Director of the National Institutes of Health, as well as from NCI, NHGRI, NHLBI, NIDA, NIMH, and NINDS. The data used for the analyses described in this manuscript were obtained from the GTEx Portal on 07/06/2023.*
